# Prevalence of vitamin D is not associated with the COVID-19 epidemic in Europe. A critical update of the existing evidence

**DOI:** 10.1101/2021.03.04.21252885

**Authors:** Dimitra Rafailia Bakaloudi, Michail Chourdakis

**Affiliations:** Laboratory of Hygiene, Social & Preventive Medicine and Medical Statistics, School of Medicine, Faculty of Health Sciences, Aristotle University of Thessaloniki, Greece

**Keywords:** Vitamin D, deficiency, COVID-19, infections, mortalities, Europe

## Abstract

**Background:** COVID-19 has emerged as a global pandemic, affecting nearly 104 million people worldwide as of February 4^th^ 2021. In previous published studies, the association between the mean Vit D status of each country and COVID-19 infection rate, and mortality among the adult population in European countries was examined. The aim of this study was to re-examine the relationship between the Vit D status of each country and COVID-19 infection, recovery, and mortality using updated data and a different methodological approach.

**Methods:** Information only form the last decade on Vit D concentration/deficiency for each country was retrieved through literature search on PubMed^®^ database. As of February, 4^th^ 2021, COVID-19 infections and mortalities per one million population as well as total recoveries were extracted from the Worldometer website. The association between vitamin D deficiency and COVID-19 infection, recovery, and mortality were explored using correlation coefficients and scatterplots.

**Findings:** The prevalence of vitamin D deficiency among European countries ranged from 6.0 (Finland) to 75.5% (Turkey), with several countries facing more than 50% of vitamin D deficiency among their population. Non-significant correlations were observed between the number of COVID-19 infections (r=0.190; p=0.374), recoveries (r_s_=0.317, p=0.131), and mortalities (r=0.129; p=0.549) per one million population, with the prevalence of vitamin D deficiency.

**Interpretation:** Prevalence of vitamin D deficiency was not significantly associated with either number of infections, recoveries or mortality rate of COVID-19 among European countries. Thus, it is an important parameter to be considered when implementing preventive measures to face COVID-19.

**Funding:** None

## INTRODUCTION

COVID-19 has become a global public health emergency, affecting more than 104 million people from 218 countries and territories^1^ in less than a year since the very first outbreak in Wuhan, China.^2^ As of February, 4^th^ 2021, the lowest and highest number of confirmed cases were reported among Oceanic (50.336) and North American (31.357.026) continents respectively.^1^ This substantial variation in the number of infections and as well as the severity and mortality of the disease can be accredited to several factors both at the state level as well as at an individual level. “State level” parameters include diverse factors, such as a country’s preparedness, actions of the governments, health infrastructure, timing of lockdown, rapid border closures, implementation of social distancing and socioeconomic status,^3^ while the “individual level” includes the sociodemographic factors and other determinants of health status such as sex, age, chronic diseases, obesity, and malnutrition.^4,5^

It is well known that malnutrition constitutes a risk factor for increased mortality and morbidity of several diseases.^6^ Protein and energy malnutrition, and other specific micronutrient deficiencies have been shown to manifest adverse effects in immunity and thereby exhibit poor prognosis of viral infections.^7^ Regarding micronutrients, the association between vitamin D (Vit D) deficiency and various diseases’ prevalence and/or severity, such as autoimmune disorders, diabetes, skeletal diseases and acute respiratory tract infections have been adequately established in the past years.^8,9^ However, evidence with regards to Vit D concentration and preventive and/or curative mechanisms of SARS-CoV-2 infection are limited^10^ or present some controversies.^11-13^ Recent studies have demonstrated the mechanisms for possible interactions between serum vitamin D concentration and rate of COVID-19 infections. Particularly, Vit D modulates the expression of angiotensin-converting enzyme 2 (ACE2), angiotensin (1-7) (Ang (1-7)), and mas receptor (MasR) axis and plays a crucial role in the protection against lung infection.^14-16^ Thereby it acts as a renin-angiotensin system (RAS) inhibitor in treating COVID-19 patients with underlying comorbidities^17,18^ and can lead to a weakening of the cytokine storm and the Acute Respiratory Syndrome (ARS) risk among COVID-19 patients, but all this evidence lacking clinical validation^19,20^.

In three recently published studies the relationship between mean concentration of Vit D and number of cases and deaths of COVID-19/1M population in 20 European countries, negative correlations were reported.^10,21,22^ In this study we aimed to re-examine the relationship between the status of Vit D and infections, recoveries, and mortalities of COVID-19 in European countries, using a bigger sample of countries and a different methodological approach.

## METHODS

### Data sources and inclusion/exclusion criteria

Information on COVID-19 infections, recoveries and mortalities, were retrieved from the Worldometer website, which provides real time statistics ^1^. This source contains data derived directly from official government reports of individual countries and/or indirectly through reliable local media resources. Data on the prevalence of Vit D deficiency among these countries were extracted by conducting a comprehensive electronic search in PubMed^®^ database (up to February 6^th^ 2021). An advanced search was performed at the level of title/abstract by using keywords such as “Vitamin D” or “25-hydroxyvitamin D_3_”, combined with “deficiency”, “prevalence” or “status” and the name of each European country. The final search string for each country and additional information about our search strategy are presented in **Supplementary Material.**

Inclusion criteria for our study were: a) Population-based studies that reported data in the last ten years; b) Studies reporting non-institutional adults (>18 years old); c) Studies defining Vit D deficiency as serum concentration <20 ng/ml or <50 nmol/l; d) Studies reporting Vit D deficiency prevalence of the sample population; e) European countries with population >1M; f) European countries in which >60.000 COVID-19 test/1M population were performed. Conference proceedings, editorials, commentaries, book chapters/book reviews and studies confined to selective sample of community-dwelling people, such as pregnant women, menopausal women and patients with diagnosed illnesses were excluded. As a last step, out of the screened articles for each country, the prevalence of Vit D deficiency data was retrieved from the most recently published study, including the most representative sample for each country.

### Data extraction

For each European country, information on COVID-19 infections, recoveries and mortalities per 1M population as of February, 4^th^ 2021, were extracted from the Worldometer website.^1^ From the selected articles reporting Vit D deficiency among these countries, name of the first author, published year, sample size, age range of the study population, mean Vit D concentration (nmol/L) and prevalence (%) of Vit D deficiency were retrieved. All data were extracted by one reviewer (DB) using a standardized excel form and were checked for accuracy by a second reviewer (MC).

### Data analysis

The relationship between the prevalence of Vit D deficiency and variables, such as number of COVID-19 infections, recoveries and mortalities per 1M population were explored with Pearson (r) or Spearman’s rank (r_s_) correlation coefficients. Scatter plots were used to visually represent the correlations. All countries were represented by a 3-letter country code according to the ISO (International Organization for Standardization) 3.166, as per the Terminology Bulletin Country Names and the Country and Region Codes for Statistical Use maintained by the United Nations Statistics Divisions.^23^ Both Pearson’s and Spearman’s correlation (2-tailed) tests were performed using the IBM SPSS^®^ version 26.0 software.

## RESULTS

A total of 24 European countries, satisfying the inclusion/exclusion criteria were selected for the analysis^24-47^ (**Table 1**). Countries excluded due to their limited population and/or lower number of COVID-19 tests were Andorra, Channel Islands, Faeroe Islands, Gibraltar, Iceland, Isle of Man, Liechtenstein Luxembourg, Malta, Monaco, Montenegro, San Marino, and Vatical city. Moreover, Albania, Belarus, Hungary, Latvia, Lithuania, Moldova, Netherlands, North Macedonia, Serbia, Slovakia, Spain, and Sweden were not part in our analysis because of the absence or the non-updated evidence regarding Vit D concentration, i.e. only data older than 10 years were available. The prevalence of Vit D deficiency ranged from 6.0-75.5% with the lowest and highest rates reported in Finland,^32^ and Turkey^45^ respectively. In 11 of the countries, the majority (>50%) of the adult population studied was Vit D deficient (i.e. <20 ng/ml or <50 nmol/l).^25,27,31,34,39-41,43,45-47^ The size of study population which was used to retrieve data for the prevalence of Vit D deficiency varied from 280 (Slovenia)^43^ to 74.235 subjects (Italy).^37^

**Table 1:** The prevalence of vitamin D deficiency among 24 European countries.

| Country | Author, Year | Sample size<br>(M/ F) | Age<br>(years) | Mean Vitamin D<br>(nmol/L) | Prevalence of<br>vitamin D<br>deficiency* (%) |
| --- | --- | --- | --- | --- | --- |
| Austria | Elmadfa et al.,<br>2017 | 541<br>(220/321) | 18-80 | No info | 48.0 |
| Belgium | Hoge et al.,<br>2015 | 697<br>(378-319) | 21-65 | 49.8 | 51.1 |
| Bosnia and<br>Herzegovina | Sokolovic et<br>al., 2016 | 1830<br>(No info) | >18 | No info | 24.3 |
| Bulgaria | Borissova et<br>al., 2013 | 2016<br>(948/1068) | 20-80 | 38.75 | 54.5 |
| Croatia | Baric et al.<br>2016 | 791<br>(131/660) | 45.5<sup>1</sup> | 54.4 | 46.1 |
| Czech Republic | Mayer et al.,<br>2012 | 560<br>(239/321) | 25-75 | 62.5 | 24.4 |
| Denmark | Cashman et al.,<br>2015 | 3409<br>(1892/1517) | 19-72 | 65.0 | 23.6 |
| Estonia | Pludowski et<br>al., 2014 | 367<br>(167/200) | 25-70 | 43.7 | 73.0 |
| Finland | Adebayo et<br>al., 2020 | 798<br>(No info) | 30-64 | 64.0 | 6.0 |
| France | Souberbielle et<br>al., 2016 | 892<br>(463/429) | 18-89 | 60.0 | 34.6 |
| Germany | Rabenberg et<br>al., 2015 | 6995<br>(3360/3635) | 18-79 | 45.6 | 61.5 |
| Greece | Dimakopoulos<br>et al., 2019 | 1084<br>(410/674) | ≥18 | 41.8 | 32.4 |
| Ireland | Cashman et<br>al., 2013 | 1118<br>(No info) | 18-84 | 56.4 | 43.7 |
| Italy | Giuliani et al.,<br>2018 | 74235<br>(18811/55424) | 18-104 | 68.5 | 33.3 |
| Norway | Petrenya et<br>al., 2020 | 4465<br>(2041/2424) | 40-69 | 64.0 | 24.7 |
| Poland | Pludowski et<br>al., 2016 | 5775<br>(1311/4464) | 16-90 | 45.0 | 65.8 |
| Portugal | Duarte et al.,<br>2020 | 3092<br>(1907/1995) | ≥18 | 42.15 | 66.6 |
| Romania | Niculescu et<br>al., 2017 | 14052<br>(1705/12347) | >21 | 49.5 | 52.0 |
| Russia | Karonova et<br>al., 2016 | 1544<br>(205/1294) | 18-75 | 54.8 | 45.7 |
| Slovenia | Hribar et al.,<br>2020 | 280<br>(128/152) | 18-74 | 50.7 | 60.7 |
| Switzerland | Sakem et al.,<br>2013 | 1291<br>(600/691) | >60 | No info | 39.2 |
| Turkey | Öztürk et al.,<br>2017 | 1161 (798/363) | 18-90 | 70.0 | 75.5 |
| United Kingdom | Aspell et al.,<br>2019 | 6004<br>(2713/3291) | >60 | 48.7 | 55.3 |
| Ukraine | Shchubelka et<br>al., 2020 | 1639 (NA) | 18-82 | 22.67 | 51.74 |

\*Serum 25-hydroxyvitamin D<sub>3</sub> concentration <20 ng/ml or <50 nmol/l; F: Female; M: Male; Mean age of the participants

As of February, 4^th^ 2021, in regard to the total number of COVID-19 infections per 1M of the total population, Russia reported the lowest with 2.684/1M population, while the Czech Republic had the highest with 94.522/1M population infections. Regarding the COVID-19 mortalities per 1M population, the lowest number, was documented in Norway (105 deaths/1M), whereas the highest number, was reported in Belgium (1.826 deaths/1M). Regarding total recoveries the lowest number was reported in Switzerland (365.4/1M) whereas in Czech Republic the highest number of recoveries was observed (84.136/1M).

Cases of COVID-19 infections per 1M population displayed a non-significant, positive correlation (r=0.190; p=0..374) with the prevalence of Vit D deficiency (**Figure 1**).

**Figure 1:**
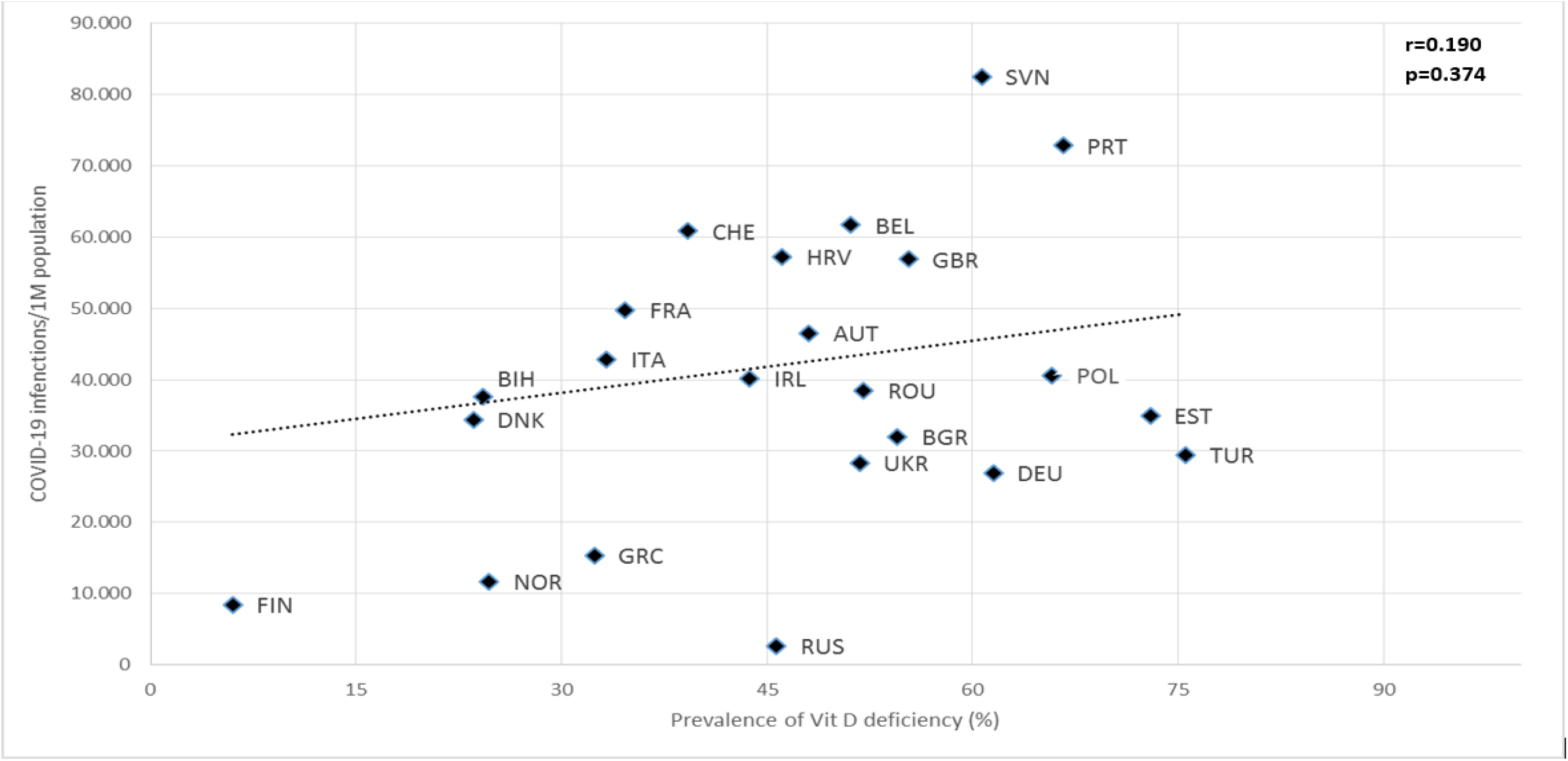
Scatter diagram of the prevalence of vitamin D deficiency against COVID-19 infections, as of February, 4^th^ 2021. AUT: Austria, BEL: Belgium, BIH: Bosnia and Herzegovina, BGR: Bulgaria, HRV: Croatia, CZE: Czech Republic, DNK: Denmark, EST: Estonia, FIN: Finland, FRA: France, DEU: Germany, GRC: Greece, IRL: Ireland, ITA: Italy, NOR: Norway, POL: Poland, PRT: Portugal, ROU: Romania, RUS: Russia, SVN: Slovenia, CHE: Switzerland, TUR: Turkey, GBR: United Kingdom, UKR: Ukraine.

As illustrated in **Figure 2**, COVID-19 mortality per 1M population was also not significantly correlated with the prevalence of Vit D deficiency (r=0.129; p=0..549).

**Figure 2:**
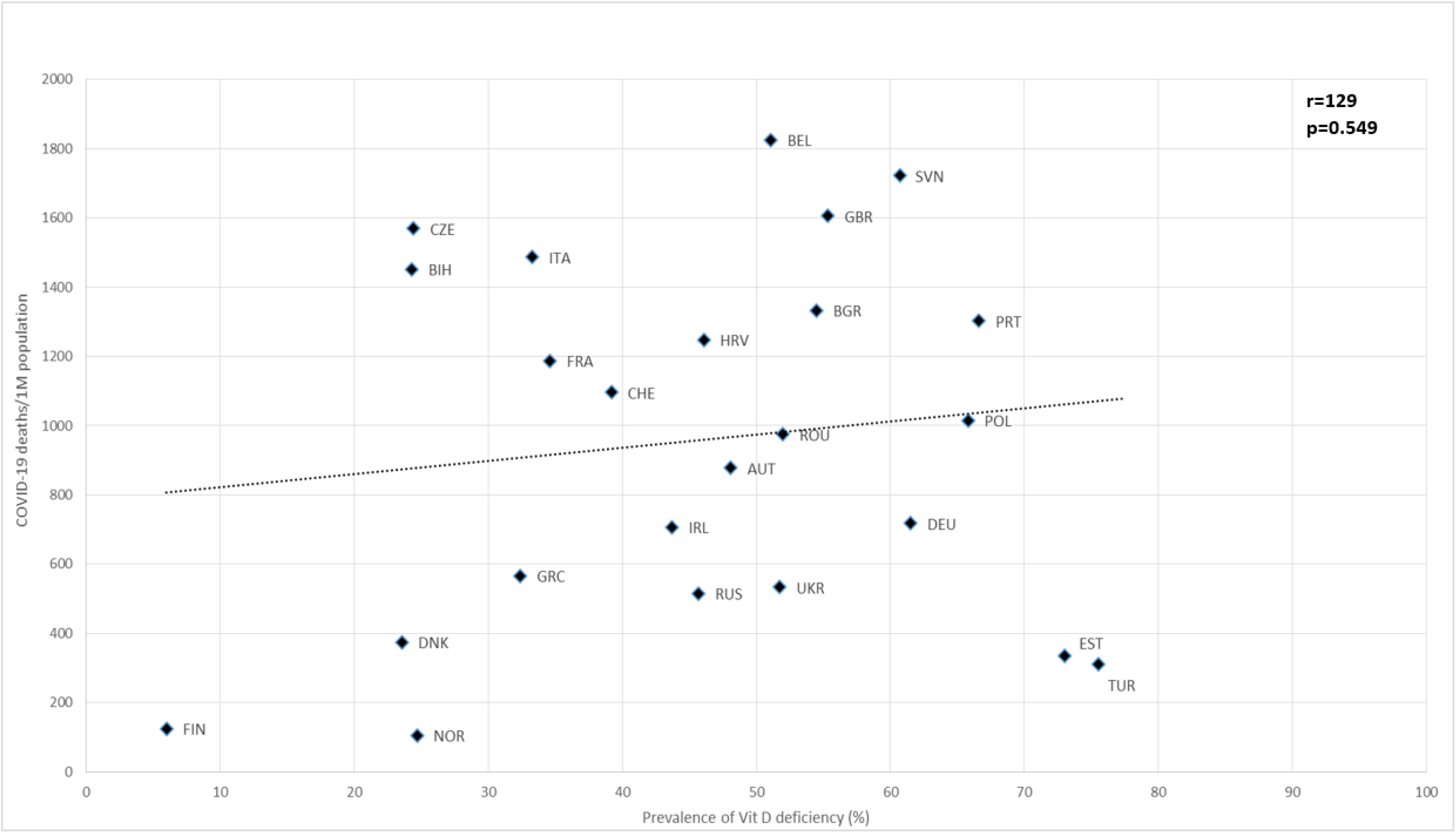
Scatter diagram of the prevalence of vitamin D deficiency against COVID-19 deaths, as of February, 4^th^ 2021. AUT Austria, BEL: Belgium, BIH: Bosnia and Herzegovina, BGR: Bulgaria, HRV: Croatia, CZE: Czech Republic, DNK: Denmark, EST: Estonia, FIN: Finland, FRA: France, DEU: Germany, GRC: Greece, IRL: Ireland, ITA: Italy, NOR: Norway, POL: Poland, PRT: Portugal, ROU: Romania, RUS: Russia, SVN: Slovenia, CHE: Switzerland, TUR: Turkey, GBR: United Kingdom, UKR: Ukraine.

As per the recovered COVID-19 cases per 1M population, similarly a non-significant correlation with the prevalence of Vit D deficiency can be observed in **Figure 3** (r_s_= 0.317, p=0.131). Moreover, the correlation and the scatter plot of prevalence of Vit D deficiency and total recoveries per COVID-19 cases per country can be found in our **Supplementary Material**

**Figure 3:**
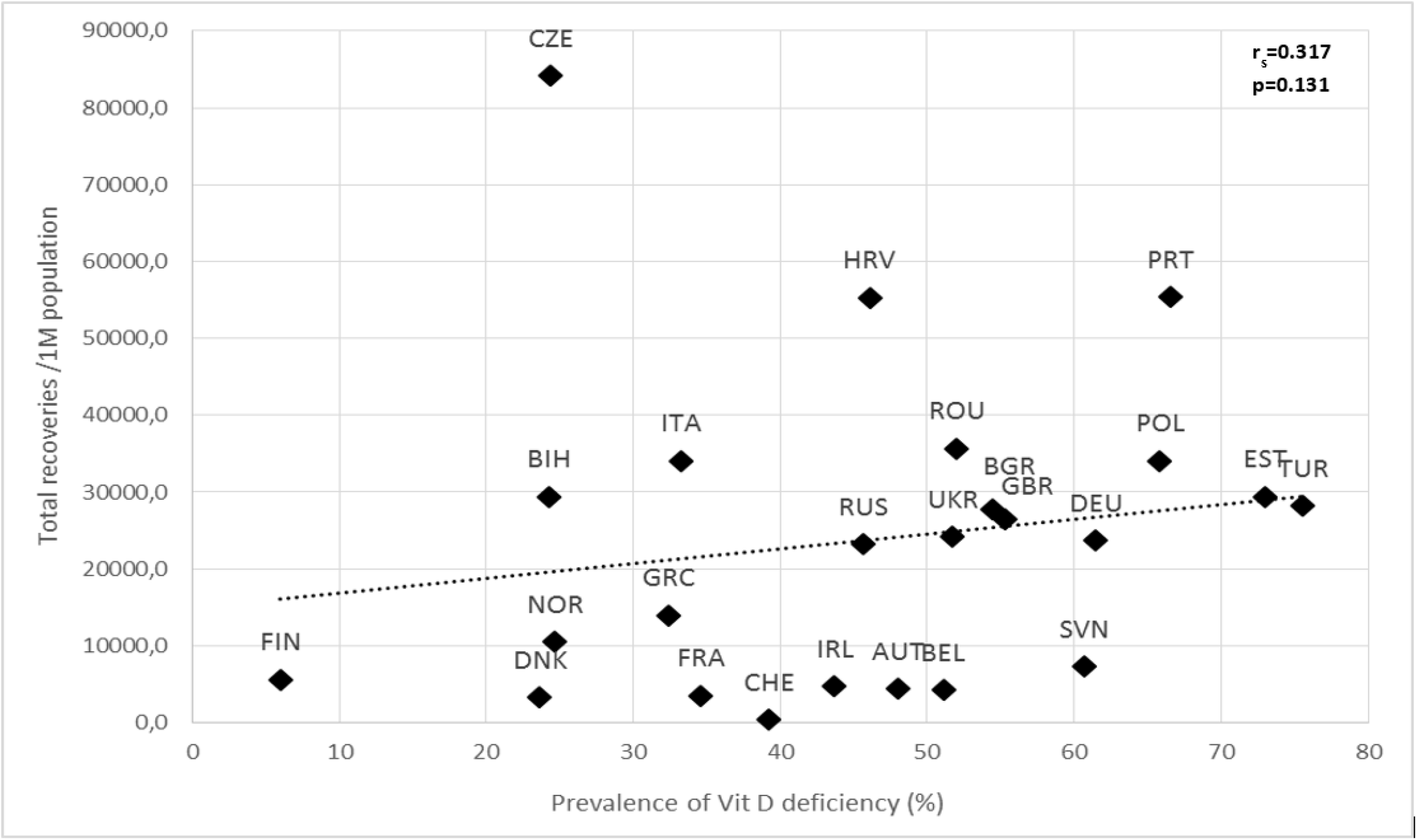
Scatter diagram of the prevalence of vitamin D deficiency against total recovered COVID-19 cases, as of February, 4^th^ 2021. AUT Austria, BEL: Belgium, BIH: Bosnia and Herzegovina, BGR: Bulgaria, HRV: Croatia, CZE: Czech Republic, DNK: Denmark, EST: Estonia, FIN: Finland, FRA: France, DEU: Germany, GRC: Greece, IRL: Ireland, ITA: Italy, NOR: Norway, POL: Poland, PRT: Portugal, ROU: Romania, RUS: Russia, SVN: Slovenia, CHE: Switzerland, TUR: Turkey, GBR: United Kingdom, UKR: Ukraine.

We also examined the relationships between COVID-19 infections, recoveries and deaths with the mean vitamin D concentration of each country, as can been seen in the **Supplementary Material**, but again no significant correlation was found.

## DISCUSSION

Our analysis concluded that the prevalence of Vit D deficiency among the European population does not constitute a strong risk factor for COVID-19 neither infection, mortality nor recovery rates. However, these findings are not in line with outcomes of similar research works published recently.^10,21,22^

According to the outcomes of our study, in several of the European countries included in this analysis, more than 50% of the adult population was Vit D deficient, which constitutes a factor that should not be disregarded in the planning of public health preventive measures. ^25,27,31,34,39-41,43,45-47^ Factors that can influence Vit D concentration include the fluctuations of sunlight exposure among seasons,^48^ especially the negligible amount of sunlight during winter and cloud covers during summer could potentially reduce the cutaneous synthesis of Vit D in some less south located countries,^49,50^ as well as extend of clothing coverage and sunscreen use.^51,52^ Furthermore, dietary sources of Vit D are limited and the obesity epidemic in the Europe, which is also related to poorer dietary choices, can worsen its deficiency.^53^ Additionally, chronic diseases such as kidney disease,^54,55^ liver disease,^56^ malignancies^57^ and genetic-epigenetic factors^58^ can influence Vit D status.

The results of our analysis showed that the prevalence of Vit D deficiency is not significantly associated with COVID-19 infections (r=0.190; p=0.374), recoveries (r_s_=0.312, p=0.138), and mortality rates (r=0.129; p=0.549). Nevertheless, the fact that our results differ from previous similar published studies^10,21,22^ can be attributed to the alternative methodological approach, which we think is correct. In our study only prevalence of Vit D deficiency for each European country was used instead of mean Vit D status for each country, a parameter which was used in those studies. A “mean” value cannot be representative of the Vit D status of a whole country because it is influenced by outliers and skewed populations. However, we have also conducted these analyses using updated data regarding Vit D mean concentration (only from the last decade) without finding any statistical significance (Supplementary Material). Therefore, we think that in the light of the most recent evidence of the COVID-19 pandemic, as well as by using updated information on Vit D deficiency prevalence for each included country, we can end up in more accurate conclusions.

Consequently, of course Vit D deficiency observed in several European countries is considered to be a crucial factor that should be frequently treated under medical supervision and not only by enhanced dietary intake^59,60^ but abuse and/or misuse of Vit D supplementations, as a method to lower the risk for COVID-19 infection is not considered appropriate.

Numerous preprints with regard to Vit D status and its association with the COVID-19 infection, recovery and mortality can be found in relevant databases (e.g., medRxiv), but these preprints have not been peer-reviewed and therefore should not be used as clinical practice guidance; additionally, such fast-track publications constitute a common risk for low quality information and should not be considered of paramount importance during the COVID-19 pandemic. An editorial in Lancet, stands equally skeptical towards findings regarding Vit D supplementation in COVID-19 patients, until more solid data become available.^61^ Moreover, also outside the scope of COVID-19, evidence regarding associations of Vit D with any outcome seems not to be convincing despite the great number of systematic reviews and meta-analyses that have been published.^62^

Although only 24 European countries satisfied our inclusion criteria, the analysis included a significant part of the European population.^63^ Therefore, the results of our study could be generalized to most of the excluded European countries too. Moreover, along with the majority of high income countries, upper-middle income countries, such as Russia and Bosnia and Herzegovina were also included in the analysis,^64^ reflecting the effect of economic status in the outcomes.

Among the limitations of our study is that the data on prevalence of Vit D deficiency of the countries included was not generated from national level surveys. Therefore, the very recently published studies (including data only from the last decade) with the most representative sample for each country’s population were carefully selected for our analysis. As described in our Methods, screened studies were limited to adults (≥18 years), as the severity of COVID-19 infections among children has been rather mild.^65^ More detailed data regarding neither COVID-19 infection rates per age nor age distribution for each country was available, and therefore correlations for these subgroups could not be performed.

Governments should implement proper preventive measures to increase awareness among the populations on the risk of Vit D deficiency rather than on its role in during the COVID-19 pandemic. Vit D supplementation should be advised only for those belonging to a high risk group of deficiency, such as new-borns, toddlers, pregnant women, elderly as well as non-Western immigrants,^66^ always under medical supervision and not as a preventive factor of COVID-19 infection. There might be several on-going randomized controlled trials (RCTs) examining the vitamin D supplementation in COVID-19 patients,^67^ however until there are solid data available from well-designed RCTs that will allow us to take relevant clinical decisions, the supplementation of Vit D as a way to prevent infection or improve recovery cannot be evidence based suggested.

## CONCLUSION

Non-significant correlation was detected between the total number of COVID-19 infections per 1M population (r=0.190; p=0.374), recoveries (r_s_= 0.317, p=0.131) and mortality per 1M population (r=0.129; p=0.549) with respect to country-specific prevalence of Vit D deficiency among 24 European countries. The different methodological approach and the updated data regarding Vit D status of each country included, led to different results to previous published studies and this should be taken into account for the clinical practice.

## Data Availability

Not applicable

## ABBREVIATIONS

1M: One million
25(OH)D: 25-hydroxyvitamin D_3_
ACE2: Angiotensin-Converting Enzyme 2
Ang (1-7): Angiotensin (1-7)
ARS: Acute Respiratory Syndrome
COVID-19: Coronavirus disease 2019
ECTS: European Calcified Tissue Society
ICU: Intensive Care Unit
ISO: International Organization for Standardization
MasR: Mas Receptor
RAS: Renin - Angiotensin System
RCT: Randomized controlled trial
RICU: Respiratory Intermediate Care Unit
SARS-CoV-2: Severe Acute Respiratory Syndrome Coronavirus
UV: Ultraviolet
UVB: Ultraviolet B
Vit D: Vitamin D

## AUTHOR CONTRIBUTIONS

Conceptualization and methodology: DB, MC

Data collection and validation: DB, MC

Data analysis: DB, MC

Writing-Original draft: DB, MC

Writing-Review and editing: DB, MC

## TABLE LEGENDS

**Supplementary Table 1:** Search terms of PubMed^®^ database.

## Supplementary figures

**Supplementary Figure 1:** Scatter diagram of the prevalence of vitamin D deficiency against total recoveries per COVID-19 cases, as of February, 4th 2021.

**Supplementary Figure 2A:** Scatter diagram of the mean vitamin D against COVID-19 infections, as of February, 4th 2021

**Supplementary Figure 2B**: Scatter diagram of the mean vitamin D against COVID-19 deaths, as of February, 4th 2021.

**Supplementary Figure 2C**: Scatter diagram of the mean vitamin D against total COVID-19 recovery cases, as of February, 4th 2021.

**Supplementary Figure 2D:** Scatter diagram of the mean vitamin D against total recoveries per COVID-19 cases, as of February, 4th 2021

