## Supplementary Material for "Prevalence of vitamin D is not associated with the COVID-19 epidemic in Europe. A critical update of the existing evidence"

Supplementary Table 1: Search terms of PubMed^®^ database.

| **Country** | **Final search string** | **No. of results** |
| --- | --- | --- |
| Albania | ("vitamin d"[Title/Abstract] OR "25 hydroxyvitamin d3"[Title/Abstract]) AND ("prevalence"[Title/Abstract] OR "deficiency"[Title/Abstract] OR "status"[Title/Abstract]) AND ("Albania"[Title/Abstract]) | 02 |
| Andorra | ("vitamin d"[Title/Abstract] OR "25 hydroxyvitamin d3"[Title/Abstract]) AND ("prevalence"[Title/Abstract] OR "deficiency"[Title/Abstract] OR "status"[Title/Abstract]) AND ("Andorra"[Title/Abstract]) | 00 |
| Austria | ("vitamin d"[Title/Abstract] OR "25 hydroxyvitamin d3"[Title/Abstract]) AND ("prevalence"[Title/Abstract] OR "deficiency"[Title/Abstract] OR "status"[Title/Abstract]) AND ("Austria"[Title/Abstract]) | 29 |
| Belarus | ("vitamin d"[Title/Abstract] OR "25 hydroxyvitamin d3"[Title/Abstract]) AND ("prevalence"[Title/Abstract] OR "deficiency"[Title/Abstract] OR "status"[Title/Abstract]) AND ("Belarus"[Title/Abstract]) | 01 |
| Belgium | ("vitamin d"[Title/Abstract] OR "25 hydroxyvitamin d3"[Title/Abstract]) AND ("prevalence"[Title/Abstract] OR "deficiency"[Title/Abstract] OR "status"[Title/Abstract]) AND ("Belgium"[Title/Abstract]) | 37 |
| Bosnia and Herzegovina | ("vitamin d"[Title/Abstract] OR "25 hydroxyvitamin d3"[Title/Abstract]) AND ("prevalence"[Title/Abstract] OR "deficiency"[Title/Abstract] OR "status"[Title/Abstract]) AND ("Bosnia and Herzegovina"[Title/Abstract]) | 01 |
| Bulgaria | ("vitamin d"[Title/Abstract] OR "25 hydroxyvitamin d3"[Title/Abstract]) AND ("prevalence"[Title/Abstract] OR "deficiency"[Title/Abstract] OR "status"[Title/Abstract]) AND ("Bulgaria"[Title/Abstract]) | 05 |
| Channel Islands | ("vitamin d"[Title/Abstract] OR "25 hydroxyvitamin d3"[Title/Abstract]) AND ("prevalence"[Title/Abstract] OR "deficiency"[Title/Abstract] OR "status"[Title/Abstract]) AND ("Channel Islands"[Title/Abstract]) | 00 |
| Croatia | ("vitamin d"[Title/Abstract] OR "25 hydroxyvitamin d3"[Title/Abstract]) AND ("prevalence"[Title/Abstract] OR "deficiency"[Title/Abstract] OR "status"[Title/Abstract]) AND ("Croatia"[Title/Abstract]) | 09 |
| Czechia | ("vitamin d"[Title/Abstract] OR "25 hydroxyvitamin d3"[Title/Abstract]) AND ("prevalence"[Title/Abstract] OR "deficiency"[Title/Abstract] OR "status"[Title/Abstract]) AND ("Czech Republic"[Title/Abstract]) | 05 |
| Denmark | ("vitamin d"[Title/Abstract] OR "25 hydroxyvitamin d3"[Title/Abstract]) AND ("prevalence"[Title/Abstract] OR "deficiency"[Title/Abstract] OR "status"[Title/Abstract]) AND ("Denmark"[Title/Abstract]) | 82 |
| Estonia | ("vitamin d"[Title/Abstract] OR "25 hydroxyvitamin d3"[Title/Abstract]) AND ("prevalence"[Title/Abstract] OR "deficiency"[Title/Abstract] OR "status"[Title/Abstract]) AND ("Estonia"[Title/Abstract]) | 04 |
| Faeroe Islands | ("vitamin d"[Title/Abstract] OR "25 hydroxyvitamin d3"[Title/Abstract]) AND ("prevalence"[Title/Abstract] OR "deficiency"[Title/Abstract] OR "status"[Title/Abstract]) AND ("Faeroe Islands"[Title/Abstract]) | 00 |
| Finland | ("vitamin d"[Title/Abstract] OR "25 hydroxyvitamin d3"[Title/Abstract]) AND ("prevalence"[Title/Abstract] OR "deficiency"[Title/Abstract] OR "status"[Title/Abstract]) AND ("Finland"[Title/Abstract]) | 103 |
| France | ("vitamin d"[Title/Abstract] OR "25 hydroxyvitamin d3"[Title/Abstract]) AND ("prevalence"[Title/Abstract] OR "deficiency"[Title/Abstract] OR "status"[Title/Abstract]) AND ("France"[Title/Abstract]) | 89 |
| Germany | ("vitamin d"[Title/Abstract] OR "25 hydroxyvitamin d3"[Title/Abstract]) AND ("prevalence"[Title/Abstract] OR "deficiency"[Title/Abstract] OR "status"[Title/Abstract]) AND ("Germany"[Title/Abstract]) | 139 |
| Gibraltar | ("vitamin d"[Title/Abstract] OR "25 hydroxyvitamin d3"[Title/Abstract]) AND ("prevalence"[Title/Abstract] OR "deficiency"[Title/Abstract] OR "status"[Title/Abstract]) AND ("Gibraltar"[Title/Abstract]) | 00 |
| Greece | ("vitamin d"[Title/Abstract] OR "25 hydroxyvitamin d3"[Title/Abstract]) AND ("prevalence"[Title/Abstract] OR "deficiency"[Title/Abstract] OR "status"[Title/Abstract]) AND ("Greece"[Title/Abstract]) | 39 |
| Hungary | ("vitamin d"[Title/Abstract] OR "25 hydroxyvitamin d3"[Title/Abstract]) AND ("prevalence"[Title/Abstract] OR "deficiency"[Title/Abstract] OR "status"[Title/Abstract]) AND ("Hungary"[Title/Abstract]) | 12 |
| Iceland | ("vitamin d"[Title/Abstract] OR "25 hydroxyvitamin d3"[Title/Abstract]) AND ("prevalence"[Title/Abstract] OR "deficiency"[Title/Abstract] OR "status"[Title/Abstract]) AND ("Iceland"[Title/Abstract]) | 23 |
| Ireland | ("vitamin d"[Title/Abstract] OR "25 hydroxyvitamin d3"[Title/Abstract]) AND ("prevalence"[Title/Abstract] OR "deficiency"[Title/Abstract] OR "status"[Title/Abstract]) AND ("Ireland"[Title/Abstract]) | 86 |
| Isle of Man | ("vitamin d"[Title/Abstract] OR "25 hydroxyvitamin d3"[Title/Abstract]) AND ("prevalence"[Title/Abstract] OR "deficiency"[Title/Abstract] OR "status"[Title/Abstract]) AND ("Isle of Man"[Title/Abstract]) | 00 |
| Italy | ("vitamin d"[Title/Abstract] OR "25 hydroxyvitamin d3"[Title/Abstract]) AND ("prevalence"[Title/Abstract] OR "deficiency"[Title/Abstract] OR "status"[Title/Abstract]) AND ("Italy"[Title/Abstract]) | 130 |
| Latvia | ("vitamin d"[Title/Abstract] OR "25 hydroxyvitamin d3"[Title/Abstract]) AND ("prevalence"[Title/Abstract] OR "deficiency"[Title/Abstract] OR "status"[Title/Abstract]) AND ("Latvia"[Title/Abstract]) | 01 |
| Liechtenstein | ("vitamin d"[Title/Abstract] OR "25 hydroxyvitamin d3"[Title/Abstract]) AND ("prevalence"[Title/Abstract] OR "deficiency"[Title/Abstract] OR "status"[Title/Abstract]) AND ("Liechtenstein"[Title/Abstract]) | 00 |
| Lithuania | ("vitamin d"[Title/Abstract] OR "25 hydroxyvitamin d3"[Title/Abstract]) AND ("prevalence"[Title/Abstract] OR "deficiency"[Title/Abstract] OR "status"[Title/Abstract]) AND ("Lithuania"[Title/Abstract]) | 04 |
| Luxembourg | ("vitamin d"[Title/Abstract] OR "25 hydroxyvitamin d3"[Title/Abstract]) AND ("prevalence"[Title/Abstract] OR "deficiency"[Title/Abstract] OR "status"[Title/Abstract]) AND ("Luxembourg"[Title/Abstract]) | 02 |
| Malta | ("vitamin d"[Title/Abstract] OR "25 hydroxyvitamin d3"[Title/Abstract]) AND ("prevalence"[Title/Abstract] OR "deficiency"[Title/Abstract] OR "status"[Title/Abstract]) AND ("Malta"[Title/Abstract]) | 00 |
| Moldova | ("vitamin d"[Title/Abstract] OR "25 hydroxyvitamin d3"[Title/Abstract]) AND ("prevalence"[Title/Abstract] OR "deficiency"[Title/Abstract] OR "status"[Title/Abstract]) AND ("Moldova"[Title/Abstract]) | 00 |
| Monaco | ("vitamin d"[Title/Abstract] OR "25 hydroxyvitamin d3"[Title/Abstract]) AND ("prevalence"[Title/Abstract] OR "deficiency"[Title/Abstract] OR "status"[Title/Abstract]) AND ("Monaco"[Title/Abstract]) | 00 |
| Montenegro | ("vitamin d"[Title/Abstract] OR "25 hydroxyvitamin d3"[Title/Abstract]) AND ("prevalence"[Title/Abstract] OR "deficiency"[Title/Abstract] OR "status"[Title/Abstract]) AND ("Montenegro"[Title/Abstract]) | 01 |
| Netherlands | ("vitamin d"[Title/Abstract] OR "25 hydroxyvitamin d3"[Title/Abstract]) AND ("prevalence"[Title/Abstract] OR "deficiency"[Title/Abstract] OR "status"[Title/Abstract]) AND ("Netherlands"[Title/Abstract]) | 104 |
| North Macedonia | ("vitamin d"[Title/Abstract] OR "25 hydroxyvitamin d3"[Title/Abstract]) AND ("prevalence"[Title/Abstract] OR "deficiency"[Title/Abstract] OR "status"[Title/Abstract]) AND ("North Macedonia"[Title/Abstract]) | 00 |
| Norway | ("vitamin d"[Title/Abstract] OR "25 hydroxyvitamin d3"[Title/Abstract]) AND ("prevalence"[Title/Abstract] OR "deficiency"[Title/Abstract] OR "status"[Title/Abstract]) AND ("Norway"[Title/Abstract]) | 116 |
| Poland | ("vitamin d"[Title/Abstract] OR "25 hydroxyvitamin d3"[Title/Abstract]) AND ("prevalence"[Title/Abstract] OR "deficiency"[Title/Abstract] OR "status"[Title/Abstract]) AND ("Poland"[Title/Abstract]) | 65 |
| Portugal | ("vitamin d"[Title/Abstract] OR "25 hydroxyvitamin d3"[Title/Abstract]) AND ("prevalence"[Title/Abstract] OR "deficiency"[Title/Abstract] OR "status"[Title/Abstract]) AND ("Portugal"[Title/Abstract]) | 16 |
| Romania | ("vitamin d"[Title/Abstract] OR "25 hydroxyvitamin d3"[Title/Abstract]) AND ("prevalence"[Title/Abstract] OR "deficiency"[Title/Abstract] OR "status"[Title/Abstract]) AND ("Romania"[Title/Abstract]) | 15 |
| Russia | ("vitamin d"[Title/Abstract] OR "25 hydroxyvitamin d3"[Title/Abstract]) AND ("prevalence"[Title/Abstract] OR "deficiency"[Title/Abstract] OR "status"[Title/Abstract]) AND ("Russia"[Title/Abstract]) | 16 |
| San Marino | ("vitamin d"[Title/Abstract] OR "25 hydroxyvitamin d3"[Title/Abstract]) AND ("prevalence"[Title/Abstract] OR "deficiency"[Title/Abstract] OR "status"[Title/Abstract]) AND ("Sun Marino"[Title/Abstract]) | 00 |
| Serbia | ("vitamin d"[Title/Abstract] OR "25 hydroxyvitamin d3"[Title/Abstract]) AND ("prevalence"[Title/Abstract] OR "deficiency"[Title/Abstract] OR "status"[Title/Abstract]) AND ("Serbia"[Title/Abstract]) | 06 |
| Slovakia | ("vitamin d"[Title/Abstract] OR "25 hydroxyvitamin d3"[Title/Abstract]) AND ("prevalence"[Title/Abstract] OR "deficiency"[Title/Abstract] OR "status"[Title/Abstract]) AND ("Slovakia"[Title/Abstract]) | 00 |
| Slovenia | ("vitamin d"[Title/Abstract] OR "25 hydroxyvitamin d3"[Title/Abstract]) AND ("prevalence"[Title/Abstract] OR "deficiency"[Title/Abstract] OR "status"[Title/Abstract]) AND ("Slovenia"[Title/Abstract]) | 05 |
| Spain | ("vitamin d"[Title/Abstract] OR "25 hydroxyvitamin d3"[Title/Abstract]) AND ("prevalence"[Title/Abstract] OR "deficiency"[Title/Abstract] OR "status"[Title/Abstract]) AND ("Spain"[Title/Abstract]) | 125 |
| Sweden | ("vitamin d"[Title/Abstract] OR "25 hydroxyvitamin d3"[Title/Abstract]) AND ("prevalence"[Title/Abstract] OR "deficiency"[Title/Abstract] OR "status"[Title/Abstract]) AND ("Sweden"[Title/Abstract]) | 113 |
| Switzerland | ("vitamin d"[Title/Abstract] OR "25 hydroxyvitamin d3"[Title/Abstract]) AND ("prevalence"[Title/Abstract] OR "deficiency"[Title/Abstract] OR "status"[Title/Abstract]) AND ("Switzerland"[Title/Abstract]) | 51 |
| Turkey | ("vitamin d"[Title/Abstract] OR "25 hydroxyvitamin d3"[Title/Abstract]) AND ("prevalence"[Title/Abstract] OR "deficiency"[Title/Abstract] OR "status"[Title/Abstract]) AND ("Turkey"[Title/Abstract]) | 98 |
| United Kingdom | ("vitamin d"[Title/Abstract] OR "25 hydroxyvitamin d3"[Title/Abstract]) AND ("prevalence"[Title/Abstract] OR "deficiency"[Title/Abstract] OR "status"[Title/Abstract]) AND ("United Kingdom"[Title/Abstract] OR "UK"[Title/Abstract]) | 360 |
| Ukraine | ("vitamin d"[Title/Abstract] OR "25 hydroxyvitamin d3"[Title/Abstract]) AND ("prevalence"[Title/Abstract] OR "deficiency"[Title/Abstract] OR "status"[Title/Abstract]) AND ("Ukraine"[Title/Abstract]) | 05 |
| Vatican City | ("vitamin d"[Title/Abstract] OR "25 hydroxyvitamin d3"[Title/Abstract]) AND ("prevalence"[Title/Abstract] OR "deficiency"[Title/Abstract] OR "status"[Title/Abstract]) AND ("Switzerland"[Title/Abstract]) | 51 |
| **Total results** | | **1950** |

** Up to February, 6^th^ 2021.

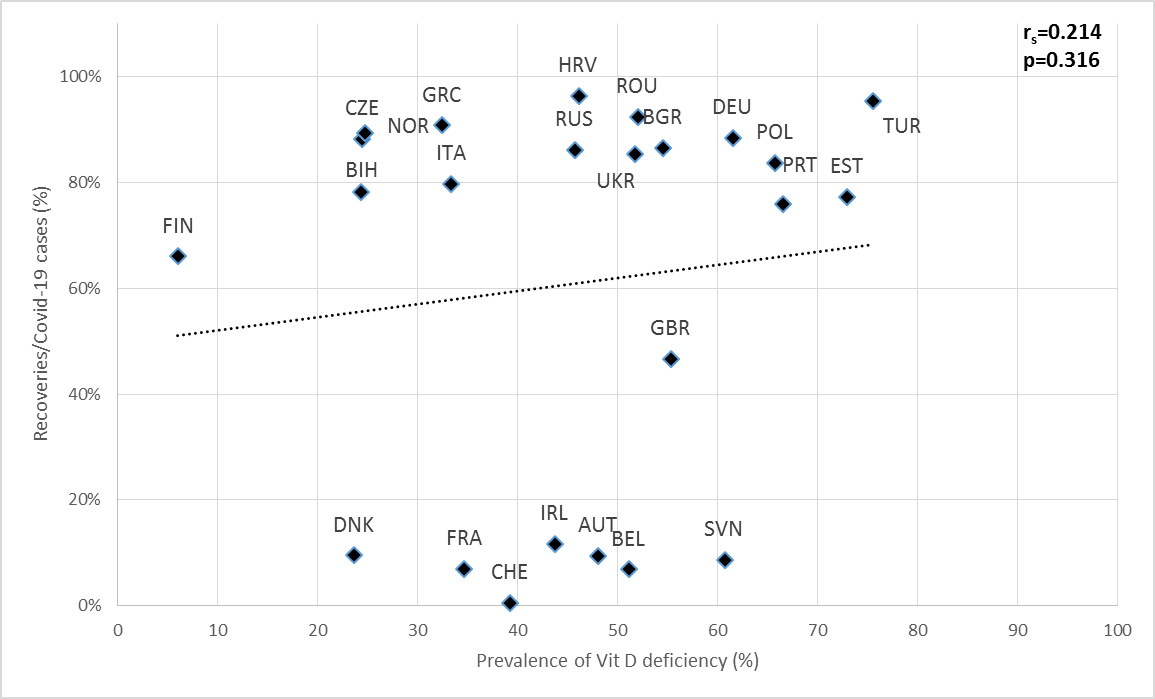

Supplementary Figure 1: Scatter diagram of the prevalence of vitamin D deficiency against total recoveries per COVID-19 cases, as of February, 4^th^ 2021.

*AUT Austria, BEL: Belgium, BIH: Bosnia and Herzegovina, BGR: Bulgaria, HRV: Croatia, CZE: Czech Republic, DNK: Denmark, EST: Estonia, FIN: Finland, FRA: France, DEU: Germany, GRC: Greece, IRL: Ireland, ITA: Italy, NOR: Norway, POL: Poland, PRT: Portugal, ROU: Romania, RUS: Russia, SVN: Slovenia, CHE: Switzerland, TUR: Turkey, GBR: United Kingdom, UKR: Ukraine.*

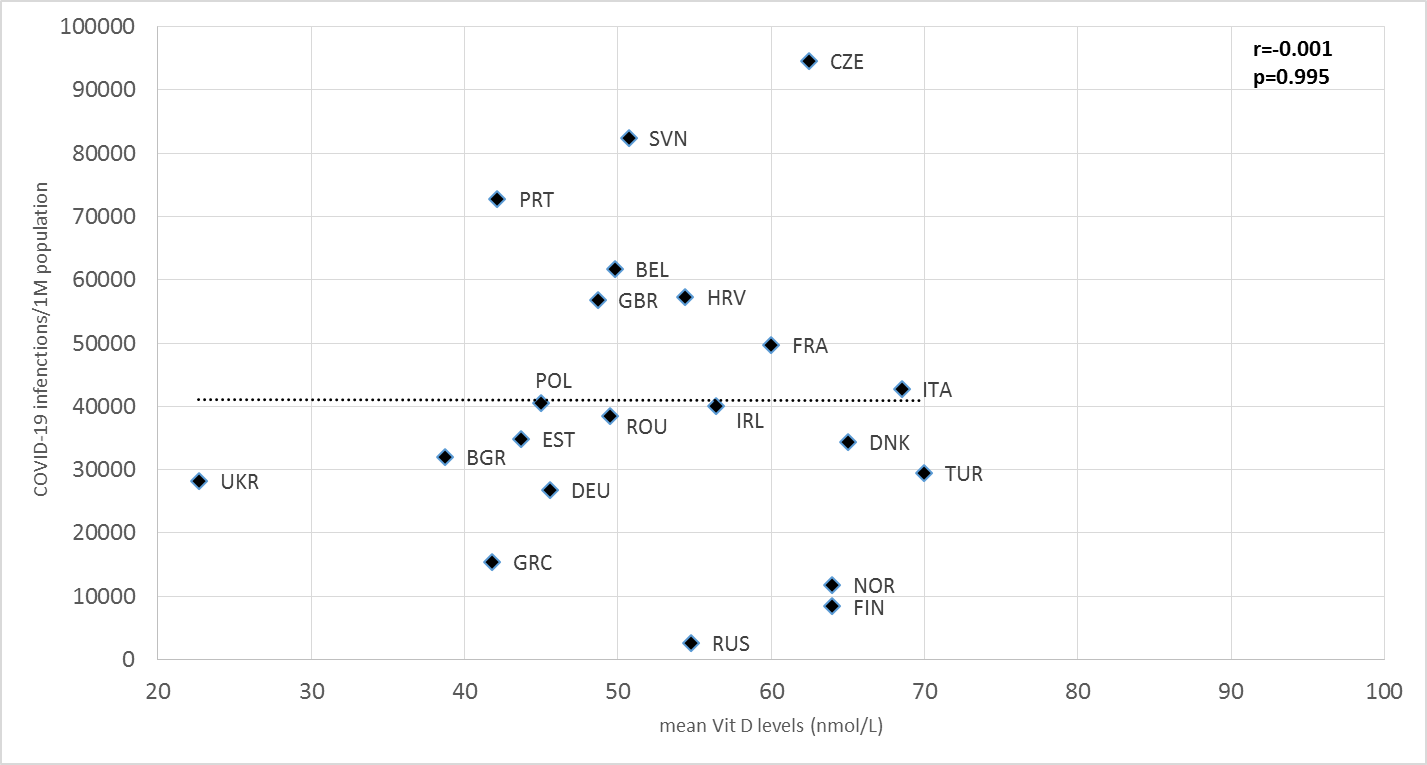

Supplementary Figure 2A: Scatter diagram of the mean vitamin D against COVID-19 infections, as of February, 4^th^ 2021.

*AUT Austria, BEL: Belgium, BIH:	Bosnia and Herzegovina, BGR: Bulgaria, HRV: Croatia, CZE: Czech Republic, DNK: Denmark, EST: Estonia, FIN: Finland, FRA: France, DEU: Germany, GRC: Greece, IRL: Ireland, ITA: Italy, NOR: Norway, POL: Poland, PRT: Portugal, ROU: Romania, RUS: Russia, SVN: Slovenia, CHE: Switzerland, TUR: Turkey, GBR: United Kingdom, UKR: Ukraine.*

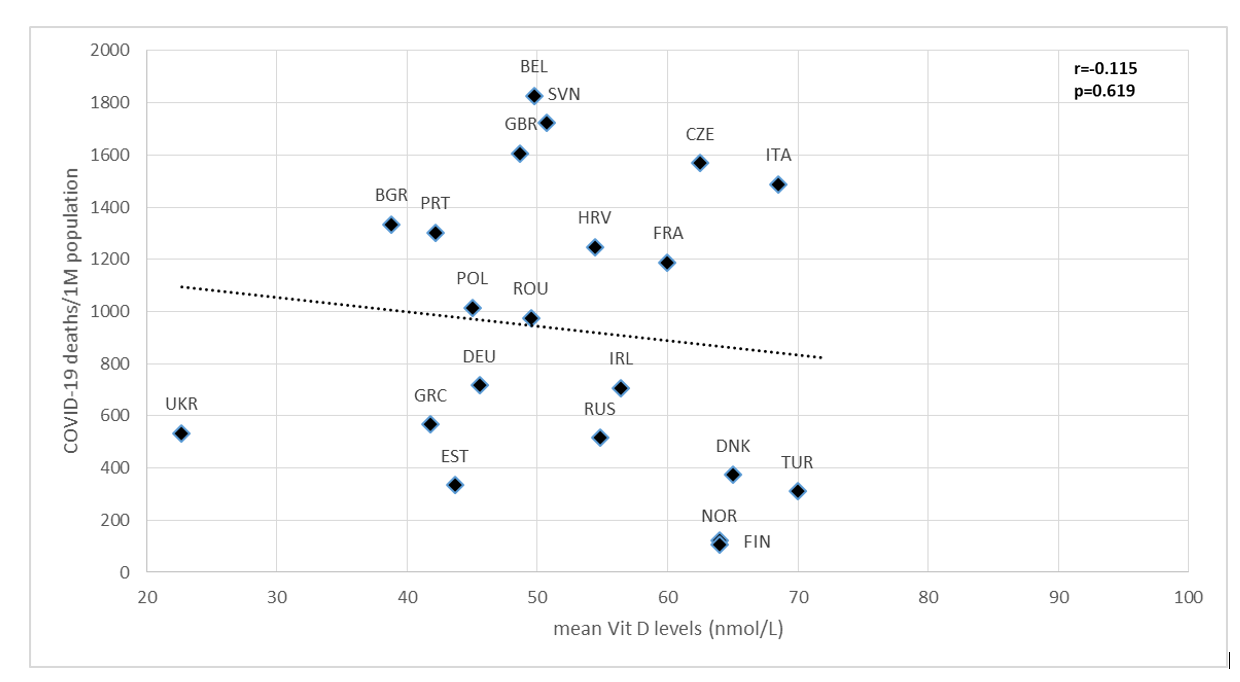

Supplementary Figure 2B: Scatter diagram of the mean vitamin D against COVID-19 deaths, as of February, 4^th^ 2021. *AUT Austria, BEL: Belgium, BIH:	Bosnia and Herzegovina, BGR: Bulgaria, HRV: Croatia, CZE: Czech Republic, DNK: Denmark, EST: Estonia, FIN: Finland, FRA: France, DEU: Germany, GRC: Greece, IRL: Ireland, ITA: Italy, NOR: Norway, POL: Poland, PRT: Portugal, ROU: Romania, RUS: Russia, SVN: Slovenia, CHE: Switzerland, TUR: Turkey, GBR: United Kingdom, UKR: Ukraine.*

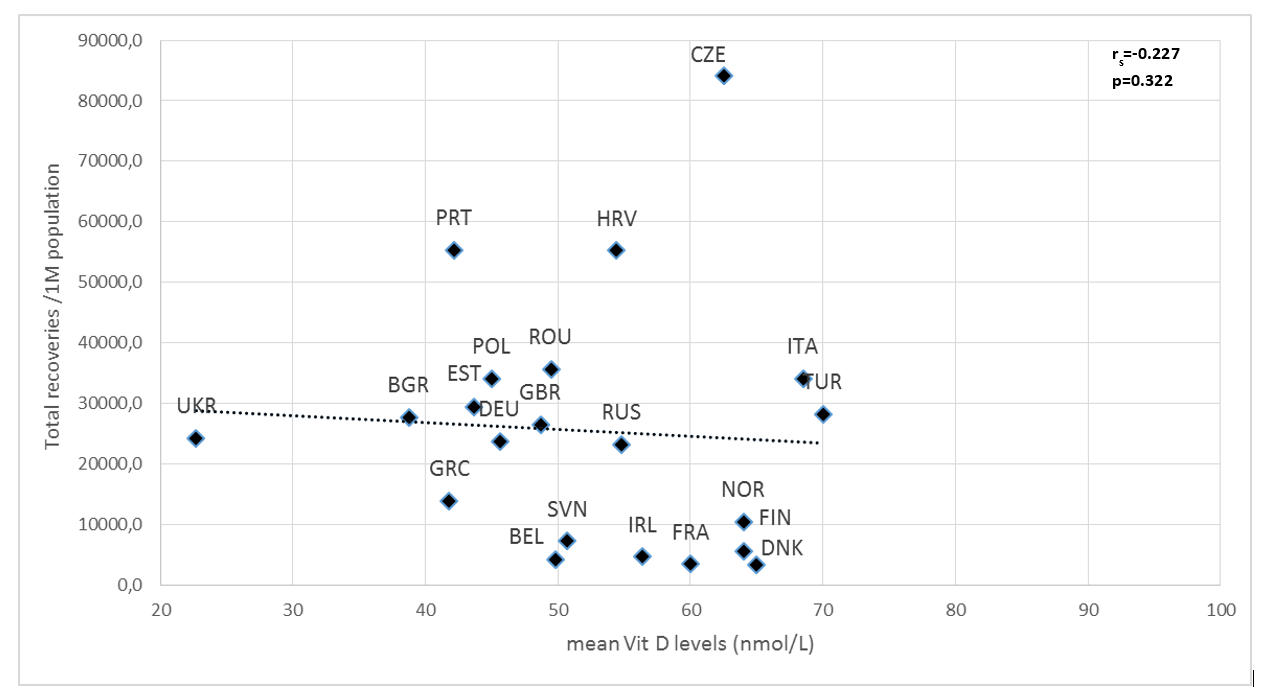

Supplementary Figure 2C: Scatter diagram of the mean vitamin D against total COVID-19 recovery cases, as of February, 4^th^ 2021.

*AUT Austria, BEL: Belgium, BIH:	Bosnia and Herzegovina, BGR: Bulgaria, HRV: Croatia, CZE: Czech Republic, DNK: Denmark, EST: Estonia, FIN: Finland, FRA: France, DEU: Germany, GRC: Greece, IRL: Ireland, ITA: Italy, NOR: Norway, POL: Poland, PRT: Portugal, ROU: Romania, RUS: Russia, SVN: Slovenia, CHE: Switzerland, TUR: Turkey, GBR: United Kingdom, UKR: Ukraine.*

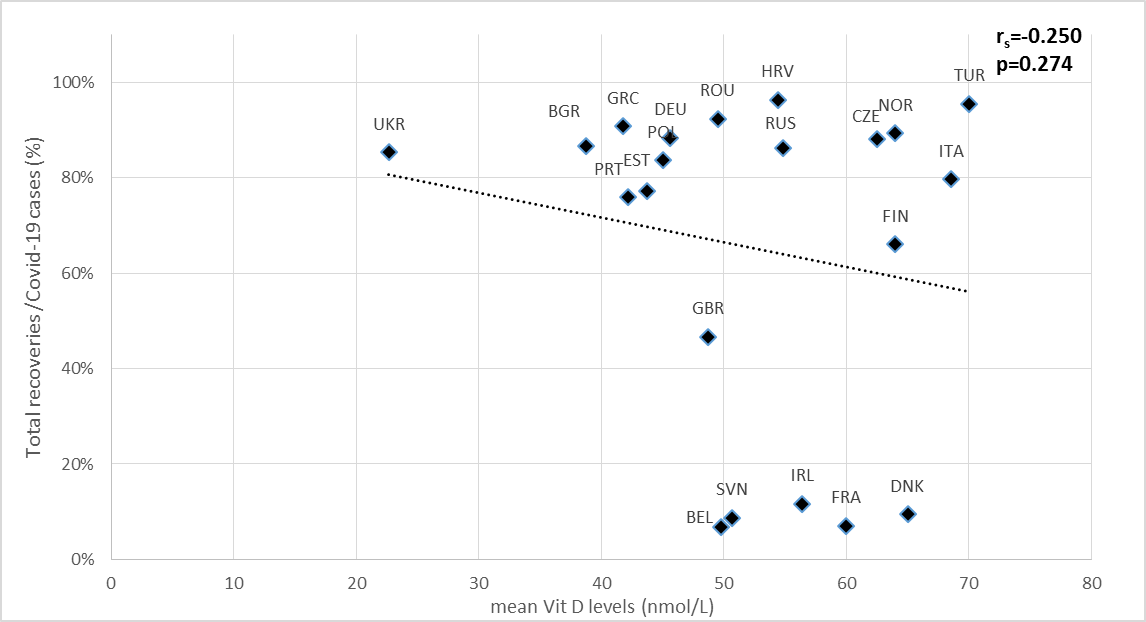

Supplementary Figure 2D: Scatter diagram of the mean vitamin D against total recoveries per COVID-19 cases, as of February, 4^th^ 2021.

*AUT Austria, BEL: Belgium, BIH:	Bosnia and Herzegovina, BGR: Bulgaria, HRV: Croatia, CZE: Czech Republic, DNK: Denmark, EST: Estonia, FIN: Finland, FRA: France, DEU: Germany, GRC: Greece, IRL: Ireland, ITA: Italy, NOR: Norway, POL: Poland, PRT: Portugal, ROU: Romania, RUS: Russia, SVN: Slovenia, CHE: Switzerland, TUR: Turkey, GBR: United Kingdom, UKR: Ukraine.*
